# Effect of vaccination on transmission of COVID-19: an observational study in healthcare workers and their households

**DOI:** 10.1101/2021.03.11.21253275

**Authors:** Anoop S V Shah, Ciara Gribben, Jennifer Bishop, Peter Hanlon, David Caldwell, Rachael Wood, Martin Reid, Jim McMenamin, David Goldberg, Diane Stockton, Sharon Hutchinson, Chris Robertson, Paul M McKeigue, Helen M Colhoun, David A McAllister

## Abstract

**Background:** The effect of vaccination for COVID-19 on onward transmission is unknown.

**Methods:** A national record linkage study determined documented COVID-19 cases and hospitalisations in unvaccinated household members of vaccinated and unvaccinated healthcare workers from 8^th^ December 2020 to 3^rd^ March 2021. The primary endpoint was COVID-19 ≥ 14 days following the first dose.

**Results:** The cohort comprised of 194,362 household members (mean age 31·1 ± 20·9 years) and 144,525 healthcare workers (mean age 44·4 ± 11·4 years). 113,253 (78·3%) of healthcare workers received at least one dose of the BNT162b2 mRNA or ChAdOx1 nCoV-19 vaccine and 36,227 (25·1%) received a second dose. There were 3,123 and 4,343 documented COVID-19 cases and 175 and 177 COVID-19 hospitalisations in household members of healthcare workers and healthcare workers respectively. Household members of vaccinated healthcare workers had a lower risk of COVID-19 case compared to household members of unvaccinated healthcare worker (rate per 100 person-years 9·40 *versus* 5·93; HR 0·70, 95% confidence interval [CI] 0·63 to 0·78). The effect size for COVID-19 hospitalisation was similar, with the confidence interval crossing the null (HR 0·77 [95% CI 0·53 to 1·10]). The rate per 100 person years was lower in vaccinated compared to unvaccinated healthcare workers for documented (20·13 *versus* 8·51; HR 0·45 [95% CI 0·42 to 0·49]) and hospitalized COVID-19 (0·97 versus 0·14; HR 0·16 [95% CI 0·09 to 0·27]). Compared to the period before the first dose, the risk of documented COVID-19 case was lower at ≥ 14 days after the second dose for household members (HR 0·46 [95% CI 0·30to 0·70]) and healthcare workers (HR 0·08 [95% CI 0·04 to 0·17]).

**Interpretation:** Vaccination of health care workers was associated with a substantial reduction in COVID-19 cases in household contacts consistent with an effect of vaccination on transmission.

## Introduction

Over 200 candidate vaccines against severe acute respiratory syndrome coronavirus 2 (SARS-CoV-2), the virus which causes coronavirus disease 2019 (COVID-19), have been developed.^1^ Several of these have shown immunogenicity in phase 2 trials,^2-4^ and three - ChAdOx1 nCoV-19, BNT162b2 mRNA and mRNA 1273 - have been shown to be effective in phase 3 trials and have been adopted for clinical use.^2,5-8^

However, while a number of studies have demonstrated that some reduction of SARS-CoV-2 transmission following vaccination may be likely,^8,9^ neither clinical trials, nor post marketing studies have compared rates of infection among close contacts of vaccinated and unvaccinated individuals. As such, the magnitude of the effect of vaccination on transmission, which is crucial for pandemic planning and informing mass-vaccination strategies,^10^ remains unknown.

Previous real-world observational studies have demonstrated the effect of vaccination on rates of infection and hospitalisation with COVID-19 outside clinical trial settings.^11-13^ However, studies examining hospitalisation did not account for occupational status, leaving the findings vulnerable to confounding by indication, since individuals in certain occupations (eg health and social care workers) are both eligible for early vaccination and at increased risk of COVID-19. Whereas the SIREN study, which did focus on a single occupational group - healthcare workers – was not large enough to examine hospitalisation.^9^ As such, the effectiveness of vaccination on rates of hospitalisation outside clinical trial settings remains uncertain.

In a nationwide study, we estimate the effect of vaccination on the risk of COVID-19 infection and hospitalisation in all healthcare workers and – as a direct measure of the effect of vaccination on transmission – all members of their households.

## Methods

### Population, data sources and record linkage

Healthcare workers were included if they were employed by the National Health Service (NHS) in Scotland on or before the 1^st^ of March 2020 (the first positive reported case of COVID-19 in Scotland) and still employed by the NHS on the 1^st^ of November 2020. Healthcare workers with a positive COVID-19 polymerase chain reaction (PCR) test before the 8^th^ December 2020 (the date when the vaccination programme was initiated) were excluded from the analysis. Data on healthcare workers and their household members were extracted and linked to multiple national datasets as previously described. In brief the Community Health Index (CHI) database, a registry of all patients registered to receive care from the NHS in Scotland (close to the complete population) was linked to the Scottish Workforce Information Standard System (SWISS) and General Practitioner Contractor Database (GPCD) databases. The CHI database was used to identify all individuals who were not themselves healthcare workers but shared a household with a healthcare worker. The healthcare worker and household data were then linked to multiple national databases including virology testing for SARS-CoV-2, general hospitalisation data, community prescribing, critical care admissions and the national register for deaths ***(Supplementary text 1 and Supplementary figure 1)***.^14^ The healthcare worker cohort was restricted to the working-age population (18-65 years of age). The household member cohort included all ages but was restricted to households with no more than one healthcare worker (4% of healthcare workers lived in multiple healthcare worker households) (***Supplementary figure 2***).

### Outcomes and exposures

Outcomes were restricted to the time period from the 8^th^ December 2020 to 3^rd^ March 2021. The pre-specified primary outcome was any positive PCR test for SARS-CoV-2 (hereafter documented COVID-19). The secondary outcome was death or hospitalisation with COVID-19 defined hospitalisations/deaths where the recorded cause was COVID-19 (ICD-10 U07·1,U07·2 or U07·5) or any hospitalisation/death occurring within 28 days of a first positive test or having first tested positive while in hospital. Where hospitalisation or death occurred without any positive test (only five and three events in healthcare workers and household members respectively), the event date was presumed to have occurred 14 days prior to the date of hospitalisation and/or death. The exposure was defined when a healthcare worker received the first dose of the BNT162b2 mRNA or ChAdOx1 nCoV-19 vaccine (the only two vaccines approved for clinical use at that time in the UK). A second dose of these vaccines was studied as a separate exposure.

### Covariates

Covariates were obtained from nationwide databases as previously described.^14^ Health board area was included as a stratifying variable, and other variables identified as potential risk factors for COVID-19 were included as covariates in regression models; age, sex, Scottish Index of Multiple Deprivation (an area-based measure of socio-economic deprivation - SIMD), ethnicity, comorbidity (as both a comorbidity count and the presence/absence of type 2 diabetes), healthcare worker role (patient facing, non-patient facing or undetermined), occupation and part-time status.

### Statistical analysis

The statistical analysis was pre-specified prior to linking the outcome and vaccination data (http://www.encepp.eu/encepp/viewResource.htm?id=39737, https://github.com/dmcalli2/hcw_vax). Person-time at risk was defined as the period from the 8th December 2020 to the date of the target event, death from a non-COVID-19 cause, or the end of follow-up (3rd March 2021), whichever came first. If household members were themselves vaccinated, person-time was censored on the day of their vaccination. Vaccination status was encoded as a time-varying categorical variable with three levels, defined with respect to the date of the first dose – unvaccinated for all days prior, intermediate for days 1 to 13, and post-dose from day 14 onwards. For additional analyses examining the effect of the second dose, we added a further level - 14 days onward from the second dose. Extended Cox regression models were used to estimate hazard ratios (HRs) for the effect of vaccination on both cases and hospitalisation, calculating robust standard errors to allow for clustering due to shared household membership and stratifying on health board area to allow for regional differences in baseline hazard.

Hazard ratios were reported unadjusted and then adjusted sequentially for socio-demographic, occupational and comorbidity covariates. Cox regression, with calendar time as the timescale, eliminates the baseline hazard rate and thus makes it unnecessary to model how the rate of COVID-19 varies over calendar time. The pre-specified primary comparison was the HR for documented cases in the period 14 days onward from the first dose compared to unvaccinated person time (i.e. before day 1 of vaccination). We also report pre-specified secondary analyses for the HRs associated with vaccination status at 7-day intervals from the date of vaccination. Analyses were performed in R Version (3·6·1).

## Results

The cohort comprised 194,362 household members (mean age 31·1±20·9) and 144,525 healthcare workers (mean age 44·4±11·4 years) (***Table 1***). Most healthcare workers were women (78·7%) whilst the majority of household members were male. Patient facing healthcare workers made up 56% of the healthcare worker population with the majority working in “front door” roles.

**Table 1:** **Baseline characteristics of vaccinated and unvaccinated healthcare workers and their households by month of first dose in the healthcare workers**

|  | Healthcare workers |  |  |  | Household members |  |  |  |
| --- | --- | --- | --- | --- | --- | --- | --- | --- |
|  | Dec-20 | Jan-21 | Feb-21 | Unvaccinated | Dec-20 | Jan-21 | Feb-21 | Unvaccinated |
| n | 49141 | 55618 | 9498 | 30268 | 65399 | 75787 | 12497 | 40679 |
| Age (mean (SD)), years | 44·92 (11·20) | 45·30 (11·22) | 46·95 (11·68) | 41·28 (11·37) | 31·29 (20·68) | 31·41 (20·95) | 32·86 (21·49) | 29·74 (20·93) |
| Sex, male (%) | 11678 (23·8) | 10961 (19·7) | 1933 (20·4) | 6175 (20·4) | 40337 (61·7) | 47347 (62·5) | 7837 (62·7) | 24875 (61·1) |
| White (%) | 47000 (95·6) | 54364 (97·7) | 9256 (97·5) | 29192 (96·4) | 62319 (95·3) | 73347 (96·8) | 11991 (96·0) | 38723 (95·2) |
| ***SIMD deprivation (%)*** |  |  |  |  |  |  |  |  |
| 1 (most deprived) | 5826 (11·9) | 8923 (16·0) | 1837 (19·3) | 5161 (17·1) | 6839 (10·5) | 10700 (14·1) | 2279 (18·2) | 6328 (15·6) |
| 2 | 8280 (16·8) | 10766 (19·4) | 2012 (21·2) | 6081 (20·1) | 10460 (16·0) | 13995 (18·5) | 2510 (20·1) | 8034 (19·7) |
| 3 | 9528 (19·4) | 11230 (20·2) | 1871 (19·7) | 5886 (19·4) | 12460 (19·1) | 15115 (19·9) | 2419 (19·4) | 7805 (19·2) |
| 4 | 11580 (23·6) | 12538 (22·5) | 2011 (21·2) | 6447 (21·3) | 16090 (24·6) | 17812 (23·5) | 2818 (22·5) | 8762 (21·5) |
| 5 (least deprived) | 13927 (28·3) | 12161 (21·9) | 1767 (18·6) | 6693 (22·1) | 19550 (29·9) | 18165 (24·0) | 2471 (19·8) | 9750 (24·0) |
|  | Dec-20 | Jan-21 | Feb-21 | Unvaccinated | Dec-20 | Jan-21 | Feb-21 | Unvaccinated |
| ***Role (%)*** |  |  |  |  |  |  |  |  |
| Patient facing |  |  |  |  |  |  |  |  |
| Front door | 19453 (39·6) | 15012 (27·0) | 2162 (22·8) | 7012 (23·2) | 26908 (41·1) | 20907 (27·6) | 2838 (22·7) | 9760 (24·0) |
| Intensive care | 1021 (2·1) | 113 (0·2) | 27 (0·3) | 90 (0·3) | 1101 (1·7) | 117 (0·2) | 19 (0·2) | 93 (0·2) |
| Other (non-AGP) | 13368 (27·2) | 12748 (22·9) | 1608 (16·9) | 5017 (16·6) | 17615 (26·9) | 17487 (23·1) | 2168 (17·3) | 6839 (16·8) |
| Other (AGP) | 1660 (3·4) | 1522 (2·7) | 146 (1·5) | 478 (1·6) | 2353 (3·6) | 2379 (3·1) | 224 (1·8) | 732 (1·8) |
| Non-patient facing | 4307 (8·8) | 11279 (20·3) | 3376 (35·5) | 11582 (38·3) | 5481 (8·4) | 15179 (20·0) | 4422 (35·4) | 15274 (37·5) |
| Undetermined | 9332 (19·0) | 14944 (26·9) | 2179 (22·9) | 6089 (20·1) | 11941 (18·3) | 19718 (26·0) | 2826 (22·6) | 7981 (19·6) |
| ***Comorbidity count %*** |  |  |  |  |  |  |  |  |
| None | 42531 (86·5) | 48330 (86·9) | 7393 (77·8) | 27284 (90·1) | 59558 (91·1) | 68798 (90·8) | 11109 (88·9) | 37226 (91·5) |
| One | 5347 (10·9) | 5920 (10·6) | 1552 (16·3) | 2467 (8·2) | 4339 (6·6) | 5166 (6·8) | 984 (7·9) | 2610 (6·4) |
| Two | 950 (1·9) | 1012 (1·8) | 379 (4·0) | 394 (1·3) | 953 (1·5) | 1111 (1·5) | 236 (1·9) | 539 (1·3) |
| Three and above | 313 (0·6) | 356 (0·6) | 174 (1·8) | 123 (0·4) | 549 (0·8) | 712 (0·9) | 168 (1·3) | 304 (0·7) |
| Type 2 diabetes | 1174 (2·4) | 1267 (2·3) | 501 (5·3) | 396 (1·3) | 1294 (2·0) | 1560 (2·1) | 350 (2·8) | 744 (1·8) |
|  | Dec-20 | Jan-21 | Feb-21 | Unvaccinated | Dec-20 | Jan-21 | Feb-21 | Unvaccinated |
| ***Vaccination data (first dose)*** |  |  |  |  |  |  |  |  |
| ChAdOx1 nCoV-19 | - | 1252 (2·3) | 3526 (37·1) | 0 (0·0) | - | 1759 (2·3) | 4340 (34·7) | 0 (0·0) |
| BNT162b2 mRNA | 48564 (98·8) | 54015 (97·1) | 5896 (62·1) | 0 (0·0) | 64636 (98·8) | 73529 (97·0) | 8038 (64·3) | 0 (0·0) |
| Unvaccinated | 0 (0·0) | 0 (0·0) | 0 (0·0) | 30268 (100·0) | 0 (0·0) | 0 (0·0) | 0 (0·0) | 40679 (100·0) |
| Unknown | 577 (1·2) | 351 (0·6) | 76 (0·8) | 0 (0·0) | 763 (1·2) | 499 (0·7) | 119 (1·0) | 0 (0·0) |
| ***Vaccination data (second dose)*** |  |  |  |  |  |  |  |  |
| ChAdOx1 nCoV-19 | 103 (0·2) | 243 (0·4) | 22 (0·2) | 0 (0·0) | 145 (0·2) | 331 (0·4) | 30 (0·2) | 0 (0·0) |
| BNT162b2 mRNA | 31983 (65·1) | 3836 (6·9) | 40 (0·4) | 0 (0·0) | 42136 (64·4) | 5307 (7·0) | 58 (0·5) | 0 (0·0) |
| Unvaccinated | 16978 (34·5) | 51482 (92·6) | 9429 (99·3) | 30268 (100·0) | 23020 (35·2) | 70055 (92·4) | 12401 (99·2) | 40679 (100·0) |
| Unknown | 77 (0·2) | 57 (0·1) | 7 (0·1) | 0 (0·0) | 98 (0·1) | 94 (0·1) | 8 (0·1) | 0 (0·0) |

**Abbreviations:** AGP – Aerosolized generating procedure; SIMD - Scottish Index of Multiple Deprivation

113,255 (78·3%) of healthcare workers received at least one dose of the BNT162b2 mRNA or ChAdOx1 nCoV-19 vaccine and 36,227 (25·1%) received a second dose. The majority of vaccinations occurred in December and January. 20·9% of healthcare workers (n=30,268 / 144,525) remained unvaccinated. 65·3% (n=32,086 / 49,141) of healthcare workers who received their first dose in December 2020 received a second dose. This dropped to 7·3% (n=4,079 / 55,618) and 0·7% (n=62 / 9,498) among those vaccinated in January and February 2021 respectively.

### Risk in household members

There were 3,123 documented COVID-19 cases and 175 hospitalisations in household members of healthcare workers. In the fully adjusted models ***(Table 2)***, for the pre-specified primary comparison (comparing the 14-day onwards post-dose period to the unvaccinated period), vaccinating the co-habiting healthcare worker was associated with a reduced risk of documented COVID-19 among household members (rate per 100 per years: 9·40 *versus* 5·93; hazard ratio [HR] 0·70, 95% confidence interval [CI] 0·63 to 0·78). Reductions of a similar magnitude were observed for COVID-19 hospitalisations (rate per 100 person-years: 0·51 *versus* 0·31; HR 0·77, 95%CI 0·53-1·10) although the wider confidence intervals included the null. In our previous report we showed that household members of patient facing healthcare workers were at a 2-fold higher risk of hospitalisation than individuals who did not share a household with a healthcare worker, i.e. at most half of all cases in household members are attributable to exposure to the health care worker and therefore preventable by preventing transmission from the health care worker.^14^ Thus, the 30% reduction which we observed for all cases is consistent with a 60% reduction in preventable cases ***(Supplementary table 1)***. If the true effect of sharing a household with a healthcare worker is lower than we previously report, the true reduction in preventable cases due to vaccination will be higher than 60%, and the converse is also true.

**Table 2:** **Effect of vaccination in healthcare workers on documented COVID-19 cases and hospitalisations in healthcare workers and their households: unvaccinated period versus period from 14-days post first dose**

|  | Healthcare workers |  |  |  | Household members |  |  |  |
| --- | --- | --- | --- | --- | --- | --- | --- | --- |
|  | Cases |  | Hospitalisations |  | Cases |  | Hospitalisations |  |
|  | Unvaccinated period | Post first dose | Unvaccinated period | Post first dose | Unvaccinated period | Post first dose | Unvaccinated | Post first dose |
| n | 144525 | 109074 | 144525 | 111081 | 194362 | 148366 | 194362 | 149689 |
| Events | 3191 | 1152 | 158 | 19 | 2037 | 1086 | 111 | 64 |
| Mean person time (days) | 40 | 45 | 41 | 45 | 41 | 45 | 41 | 45 |
| Rate per 100 person years | 20.13 | 8.51 | 0.97 | 0.14 | 9.40 | 5.93 | 0.51 | 0.35 |
| Models (Hazard ratios) |  |  |  |  |  |  |  |  |
| Unadjusted | 0.51 (0.48-0.55) |  | 0.16 (0.10-0.27) |  | 0.74 (0.67-0.82) |  | 0.83 (0.58-1.17) |  |
| Model 1 | 0.52 (0.49-0.56) |  | 0.16 (0.10-0.27) |  | 0.73 (0.66-0.81) |  | 0.81 (0.57-1.15) |  |
| Model 2 | 0.55 (0.51-0.59) |  | 0.17 (0.10-0.29) |  | 0.75 (0.68-0.83) |  | 0.86 (0.61-1.23) |  |
| Model 3 | 0.45 (0.42-0.49) |  | 0.15 (0.09-0.26) |  | 0.71 (0.63-0.78) |  | 0.77 (0.53-1.10) |  |
| Model 4 | 0.45 (0.42-0.49) |  | 0.16 (0.09-0.27) |  | 0.70 (0.63-0.78) |  | 0.77 (0.53-1.10) |  |

Results are shown for Cox regression models stratified by health board area, with the timescale being calendar time. Models were adjusted sequentially for age (using a penalized spline to allow for non-linearity) and sex (model 1); Scottish Index of Multiple Deprivation (an area-based measure of socio-economic deprivation - SIMD) and ethnicity (model 2), healthcare worker role (patient facing, non-patient facing, undetermined), occupation and part-time status (model 3); and comorbidity (as both a comorbidity count and the presence/absence of type 2 diabetes) (model 4)

In stratified analyses there was no evidence of heterogeneity in effect of vaccination by age, sex, or socio-economic deprivation of the household members or by the occupational role of the healthcare worker ***(Figure 1)***.

**Figure 1:**
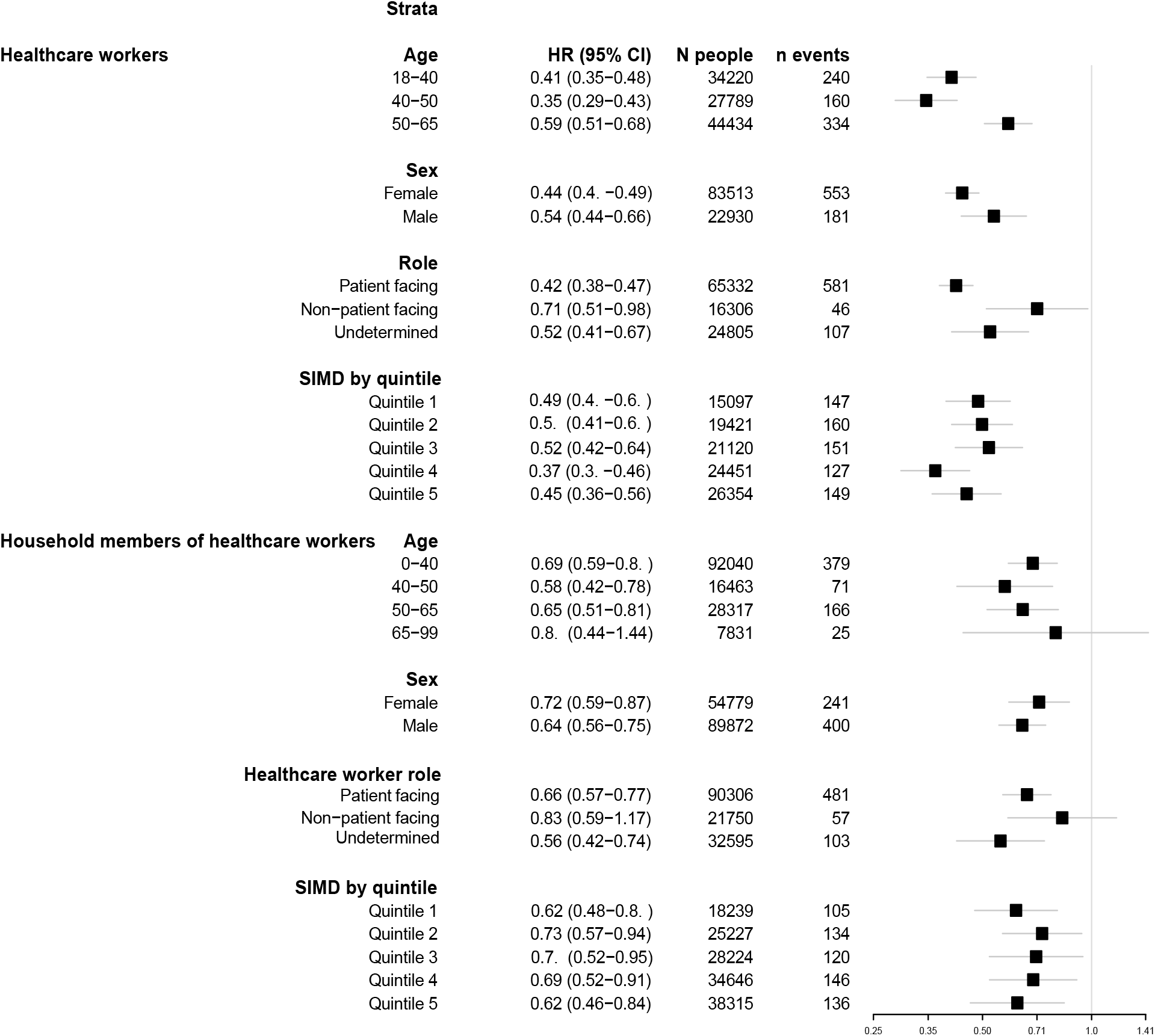
**Forest plot of adjusted hazard ratios in health care workers and their household members comparing event rate in the vaccinated period (>= 14 days following first vaccination dose in healthcare workers) to the unvaccinated period by age, sex, healthcare worker role and deprivation** Figure 1: Hazard ratios for documented COVID-19 from 14 days onwards post first dose in healthcare workers compared to unvaccinated period across healthcare workers and their household members stratified by age, sex, healthcare worker role and deprivation. Hazard ratios from from Cox models (model 4) summarised in Table 2

In the immediate period (day 1-6) following the first dose, there was a lower risk of events, but by day 7-14 there was no difference in risk associated with vaccination. Subsequent to this, vaccination was again associated with a reduced risk that became more marked with longer follow up; the associations increased in magnitude from day 14 onwards with the largest differences seen after 28 days following vaccination, HR 0·64 (95%CI 0·56-0·73) and 0·64 (95%CI 0·40-1·01) for documented cases and hospitalisations respectively, ***Table 3***).

**Table 3:** **Effect of vaccination in healthcare workers on documented COVID-19 in healthcare workers and their household at multiple time periods following the first dose**

| Population | Outcome | Unvaccinated period | Days from vaccination |  |  |  |  |
| --- | --- | --- | --- | --- | --- | --- | --- |
|  |  |  | Day 1-6 | Day 7-13 | Day 14-20 | Day 20-27 | Post day 28 |
| Healthcare workers | Cases | 1 | 0.81<br>(0.72 to 0.90) | 1.11<br>(1.02 to 1.22) | 0.54<br>(0.47 to 0.61) | 0.42<br>(0.36 to 0.49) | 0.43<br>(0.39 to 0.47) |
| Healthcare workers | Hospitalisation | 1 | 0.48<br>(0.25 to 0.92) | 0.78<br>(0.48 to 1.27) | 0.16<br>(0.06 to 0.43) | 0.13<br>(0.04 to 0.40) | 0.17<br>(0.09 to 0.32) |
| Household | Cases | 1 | 0.73<br>(0.61 to 0.87) | 1.08<br>(0.94 to 1.25) | 0.85<br>(0.73 to 0.99) | 0.68<br>(0.58 to 0.81) | 0.64<br>(0.56 to 0.73) |
| Household | Hospitalisation | 1 | 0.95<br>(0.53 to 1.69) | 1.25<br>(0.78 to 2.02) | 0.96<br>(0.56 to 1.63) | 0.83<br>(0.47 to 1.46) | 0.64<br>(0.40 to 1.01) |

Results shown are hazard ratios from Cox models adjusting for the variables shown in the footnote of Table 2 (model 4).

Compared to the unvaccinated period, the effect estimates in the fully adjusted models for the second dose (≥14 days post administration) were larger than those observed following the first dose (≥14 days post administration) for both documented cases (rate per 100 person-years 9·40 *versus* 2·98, HR 0·46 [95%CI 0·30-0·70]) and hospitalisations (rate per 100 person-years 0·51 *versus* 0·22, HR 0·68 [95%CI 0·17-2·83]), although in the case of the latter the confidence intervals were wide ***(Table 4)***.

**Table 4:**
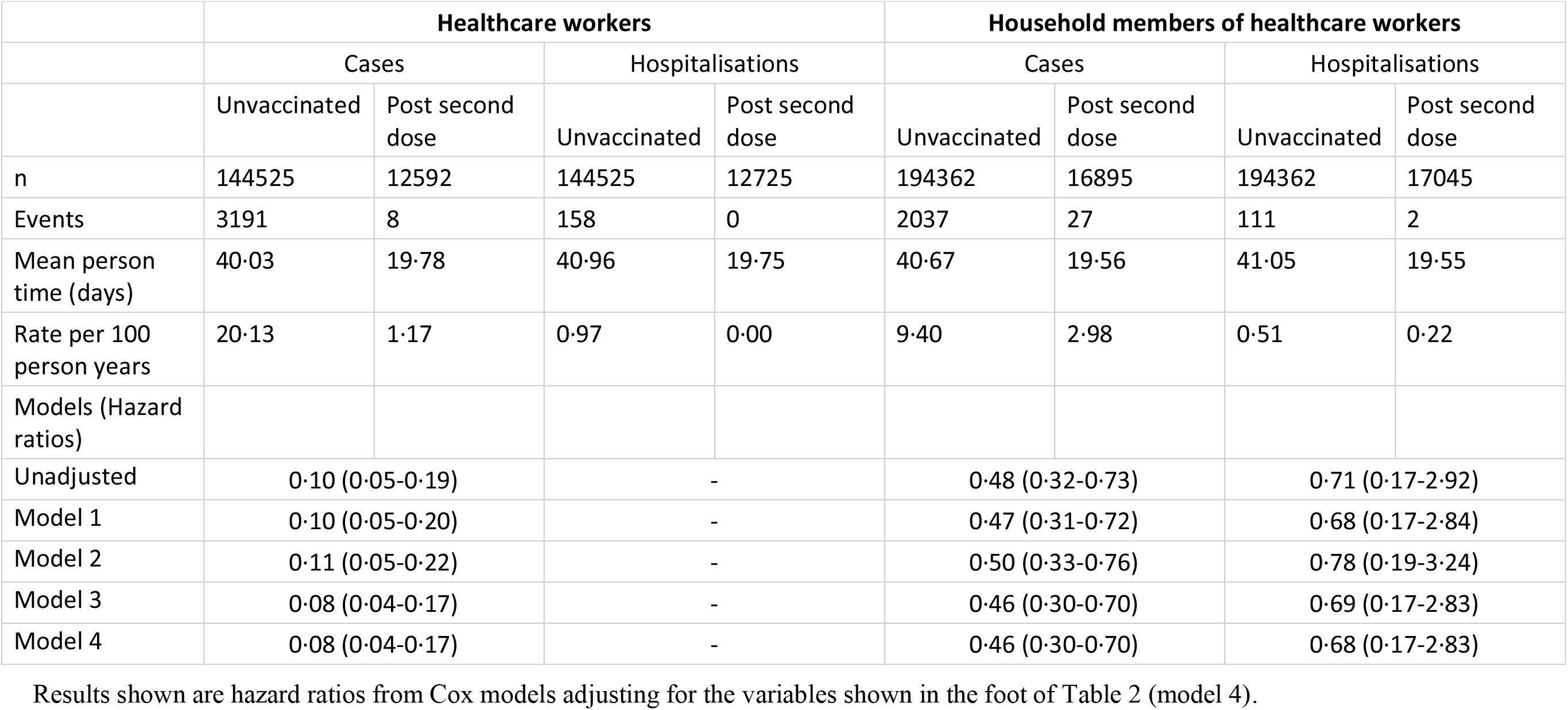
**Effect of second dose vaccination in healthcare workers on documented COVID-19 cases and hospitalisations in healthcare workers and their households: unvaccinated period versus periods from 14-days post second dose**

### Risk in healthcare workers

There were 4,343 documented cases and 177 COVID-19 hospitalisations in healthcare workers (***Table 2***). Rates of documented COVID-19 were lower in the post-dose period (≥14 days following administration) compared to the unvaccinated period (rate per 100 per years: 20·13 *versus* 8·51; HR 0·45, 95%CI 0·42-0·49, ***Table 2***). The effect on the hospitalisation rates was even larger (rate per 100 per years: 0·97 *versus* 0·14, HR 0·16, 95%CI 0·09-0·27). The strongest associations between vaccination and documented COVID-19 cases were seen in healthcare workers in patient facing roles (HR 0·42, 95%CI 0·38-0·47) ***(Figure 1)***. As for household members, the differences were larger in the later period following vaccination (***Table 3***).

Compared to the unvaccinated period, the hazard ratio for the 14-days onwards post-second dose period was larger (HR 0·08 [95%CI 0·04-0·17]; rate per 100 person years 20·13 *versus* 1·17). There were no hospitalisations in healthcare workers who received a second dose of the vaccine and as such the hazard ratio for this period could not be estimated (***Table 4***).

## Discussion

This study examined the impact of a nationwide vaccination programme on the risk of COVID-19 in healthcare workers and their household members. We found that vaccination appeared to reduce transmission. Among household members of vaccinated healthcare workers there was a 30% reduction in documented cases from day 14 post vaccination with similar, albeit not statistically significant, reductions in hospitalisation.

Consistent with previous reports,^9^ we also found a lower risk of documented cases and hospitalisation in the 1-7 day period immediately following the first dose. This occurred in both healthcare workers and their household members. Since it is too early for this to be explained by a vaccine response, it likely reflects selection bias arising from the fact that individuals with symptoms suggestive of SARS-CoV-2 infection are strongly discouraged from attending vaccination centres. This bias was expected, and partly motivated our decision - specified a priori in the statistical analysis plan - to compare rates of documented cases in the ≥14-days post first dose period to the unvaccinated period as the primary analysis.

At the time of writing a small number of post licensure studies have evaluated the impact of vaccination on symptomatic and asymptomatic COVID-19 infection.^9,11-13^ However, none of these studies examined transmission. We show an estimated reduction in risk of documented cases of 30% in household members of vaccinated healthcare workers. We observed a similar decline for hospitalisation (and a suggestion that the size of the effect continued to increase after 14-days) although this difference was not statistically significant.

Since household members of healthcare workers can also be infected via other routes, this 30% relative risk reduction is an underestimate of the ‘true’ effect of vaccination on transmission. Had all cases among household members originated from the healthcare worker with whom they lived, the 30% relative risk reduction would represent a valid estimate. However, since not all cases in a household come via the healthcare worker (based on our previous estimates the proportion is closer to half)^14^, not all cases in this group can be prevented by vaccinating healthcare workers. As such, the proportion of cases prevented by vaccinating healthcare workers which *do* arise via exposure to that healthcare worker, must be considerably larger and estimated at 60% in our analysis.

Despite a lack of previous empirical studies of transmission, this finding is biologically plausible. Vaccination reduces asymptomatic carriage of SARS-CoV-2, as has been shown in a 17,000 participants phase 3 clinical trial of the ChAdOx1 nCoV-19 vaccine, as well as in the SIREN study of healthcare workers^.8,9^ This provides a mechanistic rationale for the reduction in risk in household members, even in the absence of symptomatic infection in healthcare workers.

During our study, the Joint Committee on Vaccination and Immunisation (JCVI) in the United Kingdom changed the vaccine policy to increase the time interval between doses. Despite this, just over 25,000 healthcare workers received a second dose, which allowed us to examine the differential impact of a second dose of vaccination on transmission and infection. The relative difference in documented cases and hospitalisation in household members following the second dose was at least as large as the difference after the first dose, although for hospitalisations the numbers were small and the confidence intervals wide.

Our findings are consistent with previous studies – which did not study transmission - with respect to the direct effect on infection and hospitalisation those individuals vaccinated^.8,9^ To these studies which adjusted for a range of potential confounders, we add detailed information on occupational status. This extends previous findings since occupation is an important potential confounder as it predicts infection risk and determines access to vaccination. Our results are also consistent with the 23,324 participant SIREN study which examined risk of symptomatic and asymptomatic infections, but not hospitalisation.^9^ We add effect estimates in a homogeneous cohort with detailed occupational information which is also sufficiently large to examine the effect of vaccination on hospitalisation– confirming the large decline in both documented infection and hospitalisation in addition to the impact of vaccination on transmission.

Our existing cohort of healthcare workers and their household members provided an ideal opportunity to address the question of the effect of vaccination on transmission; transmission is commoner in households than in any other setting,^15^ healthcare workers and their household members are at increased risk of COVID-19 enriching the cohort for events,^14^ and healthcare workers have been prioritised for early-vaccination allowing this question to be addressed rapidly. Nevertheless, several points need to be considered when interpreting our results. First, the healthcare workers in our analysis were aged between 18 and 65 years of age. We are therefore unable to evaluate the impact on vaccinating elderly or vulnerable individuals. Secondly, differences in individual behaviour concerning testing before and after vaccination could introduce bias. However, this seems an unlikely explanation for our findings since similar results were seen for hospitalisation, consistent results were obtained following the first and second dose, the differences became larger over time which is biologically plausible and the direction of effect were similar in both household members and healthcare workers.

Our study has important implications for informing vaccination strategies. The JCVI in the United Kingdom recently commented on the lack of real-world evidence evaluating the role of vaccination programmes on transmission.^10^ We provide the first direct evidence that vaccinating individuals working in high-exposure settings reduces the risk to their close contacts – members of their households. This supports the specific targeting of vaccination to reduce transmission of COVID-19 in the population, such as through the vaccination of occupational groups at high risk of exposure. Similarly, clinically vulnerable groups who have been shielding during the pandemic are also more likely to include immunocompromised individuals, in whom the safety and efficacy of vaccination remains uncertain. Our findings supports the policy under consideration^16^ of vaccinating close (eg household) contacts of such individuals as a priority. Our findings further show that vaccinating healthcare workers for SARS-CoV-2 reduces but does not eliminate documented cases and hospitalisation in both those individuals vaccinated and members of their households. As such, infection prevention and control practices in healthcare settings remain of paramount importance.

Our study also provides empirical estimates of the effect of vaccination on transmission of SARS-CoV-2. These may be used to populate COVID-19 transmission models and inform decision-making on pandemic control measures.

## Conclusion

Vaccinating healthcare workers for SARS-CoV-2 reduces documented cases and hospitalisation in both those individuals vaccinated and members of their households. This is reassuring for healthcare workers, and has wide reaching implications for vaccination strategies and societal control measures.

## Data Availability

Analysis code will be made available here in an online repository on publication following peer review. Since our analysis involved data on unconsented participants, we are unable to share individual level data.

## Funding and Acknowledgements

ASVS is funded via the British Heart Foundation through an intermediate clinical research fellowship (FS/19/17/34172), and DAM is funded via a Wellcome Trust intermediate clinical fellowship and Beit fellowship (201492/Z/16/Z). The funders had no role in the study design; in the collection, analysis, and interpretation of data; in the writing of the report; and in the decision to submit the article for publication. All authors, external and internal, had full access to all of the data (including statistical reports and tables) in the study and can take responsibility for the integrity of the data and the accuracy of the data analysis. DAM thanks the Wellcome Trust for allowing him to re-deploy to Public Health Scotland to work on the response to the covid-19 pandemic.

We would like to thank Colin Tilley, Peter Ward and Morag Macpherson provided access to the SWISS database, Kathy Kenmuir, Ben Hall Lucy Munro and Susie Dodds who provided advice on re-organisations in primary and secondary care and members of the Public Health Scotland COVID-19 Health Protection Study Group and wider Public Health Scotland staff.

