## Supplement for "Effect of vaccination on transmission of COVID-19: an observational study in healthcare workers and their households"

David A McAllister^3,4^

1. Non-communicable Disease Epidemiology, London School of Hygiene and Tropical Medicine, London, UK.
2. Department of Cardiology, Imperial College NHS Trust
3. Public Health Scotland, Edinburgh, UK.
4. Institute of Health and Wellbeing, University of Glasgow, Glasgow, UK.
5. School of Health and Life Sciences, Glasgow Caledonian University, Glasgow, UK.
6. MRC Institute of Genetics & Molecular Medicine, University of Edinburgh, Edinburgh, UK.
7. Usher Institute, University of Edinburgh, Edinburgh, UK.

**Correspondence and requests for reprints:**

Dr David A McAllister

Institute of Health and Wellbeing

University of Glasgow,

Glasgow

United Kingdom

[National microbiology register (Electronic Communication of Surveillance in Scotland [ECOSS])	6](#_Toc66194438)

### Supplementary Tables and Figures

#### Supplementary table 1: Estimation of the number of cases prevented in household members by vaccinating healthcare workers as a fraction of the number of cases preventable via interrupting this route of transmission (ie those cases caused by sharing a household with a healthcare worker)

| Effect of sharing a household with a HCW | | Effect of vaccination on all detectable cases | | Effect of vaccination on preventable cases |
| --- | --- | --- | --- | --- |
| Relative risk RR_HHD_ | Attributable fraction among the exposed AF_HHD)_ *= (RR_HHD_ -1)/ RR_HHD_* | Relative risk (RR_vax_) | Relative risk reduction RRR_vax_ *= 1- RR_vax_* | Proportion of cases prevented = RRR_vax_/AF_HHD_ |
| 1.5 | 33.3% | 0.70 | 30% | 90% |
| **2.0*** | **50.0%** | **0.70** | **30%** | **60%** |
| 2.5 | 60.0% | 0.70 | 30% | 50% |
| 3.0 | 66.7% | 0.70 | 30% | 45% |

This tables illustrates shows the impact of vaccinating HCWs on transmission, taking into account that only a proportion of cases in household members could plausibly be prevented by vaccination healthcare workers, since they can also be infected via other routes.

We show the effect under different plausible estimates for the causal effect of sharing a household with a healthcare worker on the risk of developing COVID-19. The estimate in the second row (RR_HHD_ = 2.0) is close to our own previous empirical estimates for this association (BMJ 2020;371:m3582).

To perform thus calculation we made the following assumptions:-

- Vaccination of HCWs can only reduce the risk in household members by interrupting transmission from HCWs to household members and not via other mechanisms (eg by causing a modification of behavior in household members)
- That the estimated association for vaccination is a valid causal estimate – ie no bias and no confounding

#### **Supplementary figure 1: Overview of record linkage**

#### **Supplementary figure 2: Participant inclusion and exclusion healthcare workers and household members**

Died prior refers to deaths on or before the 8^th^ of December 2020

### Supplementary text 1 – Descriptions and sources of national databases used for individual record linkage

#### Scottish hospitalization record from SMR01

Comorbidities were defined from previous defined from the Scottish morbidity record 01 (SMR01) - General/Acute Inpatient & Day Case. SMR01 is an episode-based patient record relating to all inpatients and day cases discharged from non-obstetric and non-psychiatric specialties. A record is generated when a patient completes an episode of inpatient or day case care. Data collected include patient identifiable and demographic details, episode management details and general clinical information. Currently diagnoses are recorded using the ICD-10 classification and operations are recorded using the OPCS-4 classification. Further information on the national dataset and variables contained is available at <https://www.ndc.scot.nhs.uk/Data-Dictionary/SMR-Datasets//Episode-Management/SMR-Record-Type/>

#### National Records of Scotland (NRS)

The NRS covers all deaths in Scotland with approximately 55,000 deaths registered annually. The National Records of Scotland Death Records are linked with the NHS Scotland Scottish Morbidity Database which links together NHS Scotland inpatient, mental health and cancer registry datasets with the NRS Death Records. NRS records death status, cause of death and date of death.

Further information of the NRS death registry is available at <https://www.ndc.scot.nhs.uk/National-Datasets/data.asp?SubID=13>

#### Prescribing Information System (PIS)

The Prescribing Information System (PIS) is the definitive data source for all prescribing relating to all medicines and their costs that are prescribed and dispensed in the community in Scotland. The information is supplied by Practitioner & Counter Fraud Services Division (P&CFS) who is responsible for the processing and pricing of all prescriptions dispensed in Scotland. Primary care physicians write the vast majority of these prescriptions, with the remainder written by other authorised prescribers such as nurses and dentists. Also included in the dataset are prescriptions written in hospitals that are dispensed in the community. Note that prescriptions dispensed within hospitals are not included.

Further information on the Prescribing Information System operational in Scotland is available at <https://www.ndc.scot.nhs.uk/National-Datasets/data.asp?SubID=9>

#### National microbiology register (Electronic Communication of Surveillance in Scotland [ECOSS])

The Scottish microbiology surveillance registry, or ‘*Electronic Communication of Surveillance in Scotland*’ (ECOSS) as it is termed by NHS National Services Scotland, was used in the present study to provide individual patient-level data on SARS-Cov-2 testing and results. ECOSS is part of NHS Scotland’s Infection Intelligence Platform (IIP),^1,2^ which was set-up in response to the UK’s antimicrobial resistance (AMR) strategy (2013-2018) with the aim of providing “*better access to and use of surveillance data*”.^3^

Data were first collected and recorded within ECOSS in 2007. The dataset is maintained by NHS National Services Scotland on behalf of Health Protection Scotland. ECOSS is updated monthly and, as of 2017, it contained approximately 29 million records of positive microbiology laboratory specimens from across Scotland.^1^ It provides data for numerous national clinical and research activities, audit projects and Scottish Government reports, including: the identification of cases of severe infectious disease, infectious disease outbreaks and the evaluation of longer term trends in the incidence of laboratory-reported infections; surveillance of episodes of Clostridium difficile infections, Escherichia coli bacteremia, Staphylococcus aureus bacteremia and surgical site infections.^2^ NHS National Services Scotland monitors the completeness and accuracy of ECOSS data through its ‘Data Monitoring and Support Service’.^1^ Further, NHS National Services Scotland routinely informs data users of any problems affecting the accuracy or assurance of these data.

More information on the ECOSS data system is available at <https://www.hps.scot.nhs.uk/data/>

#### Scottish Workforce Information Standard System (SWISS)

The Scottish Workforce Information Standard System (SWISS) is a national human resources database held by NHS Education Scotland which contains data on all directly employed staff (ie not contracted staff such as general practitioners except where they are also directly employed in some other role) working in the NHS in Scotland. It includes data for territorial health boards, and boards providing a national service. It records the job title using a nationally agreed standard as well as, for medical and dental staff the medical specialty. It also includes data on occupation grade, part-time/whole time status, and the designated service area.

More information on the SWISS database is available at <https://turasdata.nes.nhs.scot/media/2prjxbg4/2020-06-02-workforce-report.pdf>.

#### General Practitioner Contractor Database (GPCD)

The General Practitioner Contractor Database (GPCD) includes all contracted general practitioners working in Scotland. This includes GP partners who are independent contractors, and salaried GPs, but not locum GPs. GOs in training grades are employed centrally and so are included in SWISS.

More information on the GPCD database is available at <https://www.isdscotland.org/Health-Topics/General-Practice/Workforce-and-Practice-Populations/>.

#### Rapid preliminary inpatient data (RAPID)

The RAPID database has been operational since to 2001 to monitor and predict emergency admissions and bed occupancy across National Health Service Boards in Scotland. Data from this database has already been used to provide information to NHS boards, healthcare workers and the public on the direct and indirect effects of the COVID-19 pandemic. These dashboards can be found at: <https://publichealthscotland.scot/our-areas-of-work/sharing-our-data-and-intelligence/coronavirus-covid-19-data/>

In 2015 the data collection was expanded to include data items on individual patient level data on age, sex, times of admission and discharge, ward significant facility, diagnosis and operation codes and information on patient discharge.

Further information on the RAPID databases can be found here: <https://www.ndc.scot.nhs.uk/National-Datasets/data.asp?ID=1&SubID=37>

### Supplement text 2: Definitions used to categorize healthcare workers and list of specialties, job roles and service areas by categorization

These tables are also available in machine readable formats in our public-facing github repository.

#### Definition of categorization for patient and non-patient facing and undetermined healthcare workers

| Definition | Description | Definition |
| --- | --- | --- |
| Any staff | Any member of NHS staff. | Included in SWISS or GPCD database |
| Patient facing, any | Include if likely to currently be working in patient-facing role. | Specific list of AFC roles and/or medical specialties (see below) |
| Non-patient-facing | Any member of NHS staff likely to be in a non-patient-facing role. | Specific list of AFC roles and/or medical specialties (see below) |
| Undetermined | Staff where it is not possible to allocate with confidence to patient-facing or non-patient-facing roles | Any staff not in “Patient-facing, any” or “Non-patient-facing” (see below) |

#### Definition of categorization for patient facing roles into those at front door, exposed to aerosolized generating procedures (AGP), in intensive care and other

| Definition | Description | Definition* | Comparator(s) |
| --- | --- | --- | --- |
| Patient-facing, front-door COVID19 | Include if involved in acute medical receiving of patients with possible or probable COVID19 (ie not incidental finding such as COVID19 in patient with myocardial infarction). | Specific list of AFC roles and/or medical specialties and/or designated service area (see below). | Patient-facing, other |
| Patient-facing, Intensive care | Intensive care medicine and anaesthetic specialties | Specific list of AFC roles and/or medical specialties and/or designated service area (see below). | Patient-facing, other |
| Patient-facing, Resp-oral-nasal-AGP | Include if involved in work with high risk of exposure to oral, nasal or respiratory secretions and/or aerosol generating procedures (AGP) outwith intensive care settings. | Specific list of AFC roles and/or medical specialties and/or designated service area (see below). | Patient-facing, other |
| Patient-facing, other | Patient-facing role but not front-door COVID or resp-oral-nasal-AGP or intensive care. | Patient-facing role but not front-door COVID or resp-oral-nasal-AGP or intensive care. |  |

*Nursing staff in the General Acute Nursing and Specialist Nursing and have been further assigned to specific roles according to the recorded service area. This was done for the following territorial Health Boards, in whom there is >= 95% completeness for the service area variable: - NHS Ayrshire & Arran, NHS Borders, NHS Dumfries & Galloway, NHS Forth Valley, NHS Grampian, NHS Greater Glasgow & Clyde, NHS Highland, NHS Orkney and NHS Shetland. The remaining territorial boards had lower completeness for service area (ranging from 91% to <1%)

#### Definition of medical specialties into patient facing and non-patient facing roles*

|  | Non patient-facing | Patient-facing** | | | |
| --- | --- | --- | --- | --- | --- |
|  |  | Any | Front-door | Resp-oral-nasal-AGP | Intensive care |
| Acute Internal Medicine |  | Yes | Yes |  |  |
| Allergy |  |  |  |  |  |
| Anaesthetics |  | Yes |  |  | Yes |
| Audio Vestibular Medicine |  |  |  |  |  |
| Audiological Medicine |  |  |  |  |  |
| Blood Transfusion |  |  |  |  |  |
| Breast Screening Service |  |  |  |  |  |
| Cardiology |  | Yes |  |  |  |
| Cardiothoracic Surgery |  | Yes |  |  |  |
| Chemical Pathology |  |  |  |  |  |
| Clinical Genetics |  |  |  |  |  |
| Clinical Neurophysiology |  |  |  |  |  |
| Clinical Oncology |  | Yes |  |  |  |
| Clinical Pharmacology and Therapeutics |  | Yes |  |  |  |
| Clinical Radiology |  |  |  |  |  |
| Community Psychiatry |  | Yes |  |  |  |
| Community Sexual And Reproductive Health |  | Yes |  |  |  |
| Dermatology |  |  |  |  |  |
| Diagnostic Neuropathology | Yes |  |  |  |  |
| Emergency Medicine |  | Yes | Yes |  |  |
| Endocrinology and Diabetes |  | Yes |  |  |  |
| Endodontics |  | Yes |  | Yes |  |
| Ent Surgery |  | Yes |  | Yes |  |
| Family Planning Service |  |  |  |  |  |
| Fixed & Removable Prosthodontics |  |  |  |  |  |
| Forensic Histopathology | Yes |  |  |  |  |
| Forensic Psychiatry |  |  |  |  |  |
| Gastroenterology |  | Yes |  |  |  |
| General (Internal) Medicine |  | Yes | Yes |  |  |
| General Psychiatry |  | Yes |  |  |  |
| General Surgery |  | Yes |  |  |  |
| Genito-Urinary Medicine |  | Yes |  |  |  |
| Geriatric Medicine |  | Yes | Yes |  |  |
| GP Other Than Obstetrics |  | Yes |  |  |  |
| Haematology |  |  |  |  |  |
| Histopathology | Yes |  |  |  |  |
| Homeopathy |  |  |  |  |  |
| Immunology |  |  |  |  |  |
| Infectious Diseases |  | Yes | Yes |  |  |
| Intensive Care Medicine |  | Yes |  |  | Yes |
| Medical Microbiology And Virology |  |  |  |  |  |
| Medical Oncology |  | Yes |  |  |  |
| Medical Ophthalmology |  | Yes |  |  |  |
| Microbiology |  |  |  |  |  |
| Neurology |  | Yes |  |  |  |
| Neurosurgery |  | Yes |  |  |  |
| Nuclear Medicine |  |  |  |  |  |
| Obstetrics And Gynaecology |  | Yes |  |  |  |
| Occupational Medicine |  | Yes |  |  |  |
| Old Age Psychiatry |  | Yes |  |  |  |
| Ophthalmology |  | Yes |  |  |  |
| Oral And Maxillofacial Surgery |  | Yes |  | Yes |  |
| Oral And Maxillofacial Pathology | Yes |  |  |  |  |
| Oral Medicine |  | Yes |  | Yes |  |
| Oral Microbiology |  |  |  |  |  |
| Oral Pathology |  |  |  |  |  |
| Oral Surgery |  | Yes |  | Yes |  |
| Orthodontics |  | Yes |  | Yes |  |
| Otolaryngology |  | Yes |  | Yes |  |
| Pain Management |  |  |  |  |  |
| Palliative Medicine |  |  |  |  |  |
| Plastic Surgery |  | Yes |  |  |  |
| Psychiatry Of Learning Disability |  |  |  |  |  |
| Psychotherapy |  |  |  |  |  |
| Public Health Medicine | Yes |  |  |  |  |
| Rehabilitation Medicine |  | Yes |  |  |  |
| Renal Medicine |  | Yes |  |  |  |
| Respiratory Medicine |  | Yes |  | Yes |  |
| Restorative Dentistry |  |  |  |  |  |
| Rheumatology |  | Yes |  |  |  |
| Special Care Dentistry |  | Yes |  | Yes |  |
| Surgical Dentistry |  | Yes |  | Yes |  |
| Trauma And Orthopaedic Surgery |  | Yes |  |  |  |
| Urology |  | Yes |  |  |  |
| Vascular Surgery |  | Yes |  |  |  |
| Virology |  |  |  |  |  |

There are two specialty fields in the SWISS database. These are “specialty” with around 85% completeness, and “second specialty” with lower completeness. We added data from TURAS People (which holds data on doctors in training roles) to increase the completeness to approximately 98%.

** Specialities unable to be to categorized into patient facing or non-patient facing roles were categorized into undetermined.*

***Patient facing medical specialties were categorized into front door, specialties exposed to aerosalized generating procedures and intensive care. Remaining specialties were categorized into ‘other’*

#### Definition of nursing and midwifery, allied health professionals and support services into patient facing and non-patient facing roles*

|  |  | Non patient-facing | Patient facing** | | | |
| --- | --- | --- | --- | --- | --- | --- |
| Job Family | Job Sub Family |  | Any | Front-door | Resp-oral-nasal-AGP | Intensive care |
| ADMINISTRATIVE SERVICES | FINANCE | Yes |  |  |  |  |
| ADMINISTRATIVE SERVICES | HUMAN RESOURCES | Yes |  |  |  |  |
| ADMINISTRATIVE SERVICES | INFORMATION SYSTEMS/TECHNOLOGY | Yes |  |  |  |  |
| ADMINISTRATIVE SERVICES | NA | Yes |  |  |  |  |
| ADMINISTRATIVE SERVICES | NHS24 CALL HANDLER | Yes |  |  |  |  |
| ADMINISTRATIVE SERVICES | OFFICE SERVICES | Yes |  |  |  |  |
| ADMINISTRATIVE SERVICES | PATIENT SERVICES | Yes |  |  |  |  |
| ALLIED HEALTH PROFESSION | AHP TRAINING/ADMINISTRATION | Yes |  |  |  |  |
| ALLIED HEALTH PROFESSION | AMBULANCE PARAMEDIC |  | Yes | Yes |  |  |
| ALLIED HEALTH PROFESSION | ARTS THERAPIES |  |  |  |  |  |
| ALLIED HEALTH PROFESSION | DIAGNOSTIC RADIOGRAPHY |  | Yes |  |  |  |
| ALLIED HEALTH PROFESSION | DIETETICS |  |  |  |  |  |
| ALLIED HEALTH PROFESSION | GENERIC THERAPIES |  |  |  |  |  |
| ALLIED HEALTH PROFESSION | OCCUPATIONAL THERAPY |  | Yes |  |  |  |
| ALLIED HEALTH PROFESSION | ORTHOPTICS |  |  |  |  |  |
| ALLIED HEALTH PROFESSION | ORTHOTICS |  | Yes |  |  |  |
| ALLIED HEALTH PROFESSION | PHYSIOTHERAPY |  | Yes |  | Yes |  |
| ALLIED HEALTH PROFESSION | PODIATRY |  | Yes |  |  |  |
| ALLIED HEALTH PROFESSION | PROSTHETICS |  |  |  |  |  |
| ALLIED HEALTH PROFESSION | SPEECH AND LANGUAGE THERAPY |  | Yes |  |  |  |
| ALLIED HEALTH PROFESSION | THERAPEUTIC RADIOGRAPHY |  | Yes | Yes |  |  |
| AMBULANCE SERVICES | AMBULANCE CARE ASSISTANT |  | Yes | Yes |  |  |
| AMBULANCE SERVICES | AMBULANCE TECHNICIAN |  | Yes | Yes |  |  |
| AMBULANCE SERVICES | DRIVER |  | Yes | Yes |  |  |
| AMBULANCE SERVICES | EMDC OPERATIVE | Yes |  |  |  |  |
| AMBULANCE SERVICES | OPERATIONAL MANAGER | Yes |  |  |  |  |
| AMBULANCE SERVICES | PTS DAY CONTROL | Yes |  |  |  |  |
| EMERGENCY SERVICES | AMBULANCE AUXILIARY |  | Yes | Yes |  |  |
| EMERGENCY SERVICES | AMBULANCE CARE ASSISTANT |  | Yes | Yes |  |  |
| EMERGENCY SERVICES | AMBULANCE PARAMEDIC |  | Yes | Yes |  |  |
| EMERGENCY SERVICES | AMBULANCE TECHNICIAN |  | Yes | Yes |  |  |
| EMERGENCY SERVICES | DRIVER |  | Yes | Yes |  |  |
| EMERGENCY SERVICES | EMDC OPERATIVE | Yes |  |  |  |  |
| EMERGENCY SERVICES | OPERATIONAL MANAGER | Yes |  |  |  |  |
| EMERGENCY SERVICES | PTS DAY CONTROL |  |  |  |  |  |
| HEALTHCARE SCIENCES | BIOMEDICAL SCIENCES LIFE |  |  |  |  |  |
| HEALTHCARE SCIENCES | CLIN PHOTO/ILLUSTRATE PHYSICAL |  |  |  |  |  |
| HEALTHCARE SCIENCES | CLINICAL PERFUSION PHYSIOLOGY |  |  |  |  |  |
| HEALTHCARE SCIENCES | CLINICAL PHYSIOLOGY |  |  |  |  |  |
| HEALTHCARE SCIENCES | CLINICAL SCIENCES LIFE |  |  |  |  |  |
| HEALTHCARE SCIENCES | CLINICAL SCIENCES PHYSICAL |  |  |  |  |  |
| HEALTHCARE SCIENCES | CLINICAL SCIENCES PHYSIOLOGY |  |  |  |  |  |
| HEALTHCARE SCIENCES | CLINICAL TECHNOLOGY LIFE |  |  |  |  |  |
| HEALTHCARE SCIENCES | CLINICAL TECHNOLOGY PHYSICAL |  |  |  |  |  |
| HEALTHCARE SCIENCES | MAXILLOFACIAL PROS PHYSICAL |  |  |  |  |  |
| HEALTHCARE SCIENCES | NA |  |  |  |  |  |
| HEALTHCARE SCIENCES | STERILE SERVICES LIFE |  |  |  |  |  |
| MEDICAL AND DENTAL SUPPORT | DENTAL NURSING |  | Yes |  | Yes |  |
| MEDICAL AND DENTAL SUPPORT | DENTAL TECHNOLOGY |  | Yes |  | Yes |  |
| MEDICAL AND DENTAL SUPPORT | OPERATING DEPARTMENT |  | Yes |  |  |  |
| MEDICAL AND DENTAL SUPPORT | ORAL HEALTH |  | Yes |  | Yes |  |
| MEDICAL AND DENTAL SUPPORT | PHYSICIANS ASSISTANT |  |  |  |  |  |
| MEDICAL AND DENTAL SUPPORT | THEATRE SERVICES |  | Yes |  |  |  |
| MEDICAL SUPPORT | OPERATING DEPARTMENT |  | Yes |  |  |  |
| MEDICAL SUPPORT | PHYSICIANS ASSISTANT |  | Yes |  |  |  |
| MEDICAL SUPPORT | THEATRE SERVICES |  | Yes |  |  |  |
| NURSING AND MIDWIFERY | COMMUNITY CHILDREN'S NURSING |  |  |  |  |  |
| NURSING AND MIDWIFERY | MIDWIFERY DIRECT CC |  | Yes |  |  |  |
| NURSING AND MIDWIFERY | MIDWIFERY INDIRECT CC |  | Yes |  |  |  |
| NURSING AND MIDWIFERY | NA |  |  |  |  |  |
| NURSING AND MIDWIFERY | NEONATAL MIDWIFERY CC |  | Yes |  |  |  |
| NURSING AND MIDWIFERY | NEONATAL MIDWIFERY DIRECT CC |  | Yes |  |  |  |
| NURSING AND MIDWIFERY | NEONATAL MIDWIFERY INDIRECT CC |  | Yes |  |  |  |
| NURSING AND MIDWIFERY | NEONATAL NURSING DIRECT CC |  | Yes |  |  |  |
| NURSING AND MIDWIFERY | NEONATAL NURSING INDIRECT CC |  | Yes |  |  |  |
| NURSING AND MIDWIFERY | NHS 24 NURSING | Yes |  |  |  |  |
| NURSING AND MIDWIFERY | NURSING TRAINING/ADMIN/MGT |  |  |  |  |  |
| NURSING AND MIDWIFERY | PAEDIATRIC NURSING |  | Yes | Yes |  |  |
| NURSING AND MIDWIFERY | PRACTICE NURSING |  | Yes |  |  |  |
| NURSING AND MIDWIFERY | PUBLIC HEALTH NURSING |  |  |  |  |  |
| NURSING AND MIDWIFERY | SCHOOL NURSING |  |  |  |  |  |
| NURSING AND MIDWIFERY | SEXUAL AND REPRODUCTIVE HEALTH |  |  |  |  |  |
| NURSING AND MIDWIFERY | SPECIALIST NURSING |  | Yes |  |  |  |
| NURSING AND MIDWIFERY | STAFF NURSERY |  |  |  |  |  |
| NURSING AND MIDWIFERY | TREATMENT ROOM NURSING |  | Yes |  |  |  |
| NURSING AND MIDWIFERY | BANK NURSING |  | Yes |  |  |  |
| NURSING AND MIDWIFERY | BTS NURSING |  | Yes | Yes |  |  |
| NURSING AND MIDWIFERY | CARE OF THE ELDERLY NURSING |  | Yes | Yes |  |  |
| NURSING AND MIDWIFERY | COMMUNITY GENERAL NURSING |  | Yes |  |  |  |
| NURSING AND MIDWIFERY | DISTRICT NURSING |  | Yes |  |  |  |
| NURSING AND MIDWIFERY | FAMILY PLANNING NURSING |  |  |  |  |  |
| NURSING AND MIDWIFERY | GENERAL ACUTE NURSING |  | Yes | Yes |  |  |
| NURSING AND MIDWIFERY | HEALTH VISITOR NURSING |  |  |  |  |  |
| NURSING AND MIDWIFERY | LEARNING DISABILITIES NURSING |  | Yes |  |  |  |
| NURSING AND MIDWIFERY | MENTAL HEALTH NURSING |  | Yes |  |  |  |
| NURSING AND MIDWIFERY | MIDWIFERY |  | Yes |  |  |  |
| OTHER THERAPEUTIC | GENETIC COUNSELLING |  |  |  |  |  |
| OTHER THERAPEUTIC | NA |  |  |  |  |  |
| OTHER THERAPEUTIC | OPTOMETRY |  |  |  |  |  |
| OTHER THERAPEUTIC | PHARMACY |  |  |  |  |  |
| OTHER THERAPEUTIC | PHARMACY TECHNICIANS | Yes |  |  |  |  |
| OTHER THERAPEUTIC | PLAY SPECIALIST |  | Yes |  |  |  |
| OTHER THERAPEUTIC | PSYCHOLOGY |  |  |  |  |  |
| PERSONAL AND SOCIAL CARE | CARE AT HOME |  | Yes |  |  |  |
| PERSONAL AND SOCIAL CARE | HEALTH PROMOTION |  |  |  |  |  |
| PERSONAL AND SOCIAL CARE | HOSPITAL CHAPLAINCY |  |  |  |  |  |
| PERSONAL AND SOCIAL CARE | RESIDENTIAL / DAY CARE |  | Yes |  |  |  |
| PERSONAL AND SOCIAL CARE | SOCIAL WORK |  |  |  |  |  |
| SENIOR MANAGERS | NA | Yes |  |  |  |  |
| SUPPORT SERVICES | CATERING SERVICES |  |  |  |  |  |
| SUPPORT SERVICES | DOMESTIC SERVICES |  |  |  |  |  |
| SUPPORT SERVICES | ESTATES |  |  |  |  |  |
| SUPPORT SERVICES | GENERAL SERVICES |  |  |  |  |  |
| SUPPORT SERVICES | GROUNDS SERVICES |  |  |  |  |  |
| SUPPORT SERVICES | HOTEL SERVICES |  |  |  |  |  |
| SUPPORT SERVICES | LAUNDRY/LINEN SERVICES |  |  |  |  |  |
| SUPPORT SERVICES | NA |  |  |  |  |  |
| SUPPORT SERVICES | PORTERING SERVICES |  | Yes |  |  |  |
| SUPPORT SERVICES | SECURITY SERVICES |  |  |  |  |  |
| SUPPORT SERVICES | STERILE SERVICES |  |  |  |  |  |
| SUPPORT SERVICES | STORES SERVICES |  |  |  |  |  |
| SUPPORT SERVICES | TRANSPORT SERVICES |  |  |  |  |  |
| UNALLOCATED / NOT KNOWN | NA |  |  |  |  |  |
| UNALLOCATED / NOT KNOWN | NOT KNOWN |  |  |  |  |  |

** Roles unable to be to categorized into patient facing or non-patient facing were categorized into undetermined.*

***Patient facing roles were categorized into front door, specialties exposed to aerosalized generating procedures and intensive care. Remaining specialties were categorized into ‘other’*

#### Definition of relevant service areas

|  | Front –door | Respiratory-oro-aerosol –generating procedures | Intensive care |
| --- | --- | --- | --- |
| Accident and Emergency | Yes |  |  |
| Anaesthetics |  |  | Yes |
| Ear Nose & Throat |  | Yes |  |
| Endocrinology & Diabetes |  |  |  |
| Gastroenterology |  |  |  |
| General Medicine | Yes |  |  |
| Infectious Diseases | Yes |  |  |
| Intensive Care |  |  | Yes |
| Neonatal/SCBU |  |  | Yes |
| Oral & Maxillofacial |  | Yes |  |
| Respiratory |  | Yes |  |
| Restorative Dentistry |  | Yes |  |
| Rheumatology |  |  |  |
| Surgical Dentistry |  | Yes |  |

### Supplement text 3: Comorbidity definitions

The following ICD-10 definitions were used to define each comorbidity (British National Formulary definitions are included in our public github repository).

| CIRCULATORY DISORDERS | |
| --- | --- |
| Ischaemic heart disease | I20-I25 |
| Other heart disease | I00-102 |
|  | I05-I09 |
|  | I10-I15 |
|  | I26-I28 |
|  | I30-I52 |
| Cerebrovascular Disease | I60-I69 |
|  | G45 = Transient cerebral ischaemic attacks |
|  | G46 = Vascular syndromes of brain in cerebrovascular diseases |
| Other circulatory system diseases | I70-I79 Diseases of arteries, arterioles and capillaries |
|  | I80-I89 Diseases of veins, lymphatic vessels and lymph nodes, not elsewhere classified |
|  | I95-I99 Other and unspecified disorders of the circulatory system |
|  | Z958 presence of vascular implants and grafts |
|  | Z959 presence of vascular implants and grafts NOS |
| Neurological diseases except inflammatory | |
| Epilepsy | G40-G47 Episodic and paroxysmal disorders |
| Mono and polyneuropathies | G50-G59 Nerve, nerve root and plexus disorders |
|  | G60-G64 Polyneuropathies and other disorders of the peripheral nervous system |
| Other neurological conditions | G10-G14 Systemic atrophies primarily affecting the central nervous system |
|  | G20-G26 Extrapyramidal and movement disorders |
|  | G30-G32 Other degenerative diseases of the nervous system |
|  | G35-G37 Demyelinating diseases of the central nervous system |
|  | G70-G73 Diseases of myoneural junction and muscle |
|  | G80-G83 Cerebral palsy and other paralytic syndromes |
|  | G90-G99 Other disorders of the nervous system |
| Respiratory diseases | |
| Acute respiratory infections | J00-J06 Acute upper respiratory infections |
|  | J09-J18 Influenza and pneumonia |
|  | J20-J22 Other acute lower respiratory infections |
| Asthma | J45 Asthma |
|  | J46 Status asthmaticus |
| Other Chronic lower respiratory disease | J40 Bronchitis, not specified as acute or chronic |
|  | J41 Simple and mucopurulent chronic bronchitis |
|  | J42 Unspecified chronic bronchitis |
|  | J43 Emphysema |
|  | J44 Other chronic obstructive pulmonary disease |
|  | J47 Bronchiectasis |
|  | J60-J70 Lung diseases due to external agents |
|  | J80-J84 Other respiratory diseases principally affecting the interstitium |
|  | J85-J86 Suppurative and necrotic conditions of lower respiratory tract |
|  | J90-J94 Other diseases of pleura |
|  | J95-J99 Other diseases of the respiratory system |
|  | G473 Sleep apnoea |
| Tuberculosis | A15 Respiratory tuberculosis, bacteriologically and histologically confirmed |
|  | A16 Respiratory tuberculosis, not confirmed bacteriologically or histologically |
|  | A17 Tuberculosis of nervous system |
|  | A18 Tuberculosis of other organs |
|  | A19 Miliary tuberculosis |
| Connective tissue diseases | |
| Connective tissue disorder | M050 Felty syndrome |
|  | M051 Rheumatoid lung disease |
|  | M052 Rheumatoid vasculitis |
|  | M053 Rheumatoid arthritis with involvement of other organs and systems |
|  | M058 Other seropositive rheumatoid arthritis |
|  | M059 Seropositive rheumatoid arthritis, unspecified |
|  | M060 Seronegative rheumatoid arthritis |
|  | M063 Rheumatoid nodule |
|  | M069 Rheumatoid arthritis, unspecified |
|  | M32 Systemic lupus erythematosus |
|  | M332 Polymyositis |
|  | M353 Polymyalgia rheumatica |
|  | M34 Systemic sclerosis |
| Decompensated liver disease | C22.0 hepatocellular carcinoma |
|  | I85.0 Oesophageal varices |
|  | I98.3 Oesophageal varices |
|  | K70.4 Alcoholic hepatic failure |
|  | K72.0 Acute and subacute failure of the liver |
|  | K72.1 Chronic hepatic failure |
|  | K72.9 Hepatic coma |
|  | K76.7 hepatorenal syndrome |
|  | R18 Ascites |
| Kidney disease | |
| Advanced Chronic kidney disease or RRT | N18.3 Chronic kidney disease stage 3 |
|  | N18.4 Chronic kidney disease stage 4 |
|  | N18.5 Chronic kidney disease stage 5 |
|  | Z490 Care involving dialysis |
|  | Z491 Care involving dialysis |
|  | Z492 Care involving dialysis |
|  | Z940 Kidney transplant status |
|  | Z992 Dependence on renal dialysis |
| Other chronic kidney disease | N00-N08 Glomerular diseases |
|  | N10-N16 Renal Tubulo-interstitial disease |
|  | N18.1 Chronic kidney disease stage 1 |
|  | N18.2 Chronic kidney disease stage 2 |
|  | N18.9 Chronic kidney disease unspecified |
| Diabetes | |
|  | E10 Type 1 diabetes mellitus |
|  | E11 Type 2 diabetes mellitus |
|  | E12 Malnutrition-related diabetes mellitus |
|  | E13 Other specified diabetes mellitus |
|  | E14 Unspecified diabetes mellitus |
| Malignant neoplasms | C00-C97 |
| Lung cancers | C34 Malignant neoplasm  of bronchus and lung |
| Blood cancers | C81-C96 Malignant neoplasms, stated or presumed to be primary, of lymphoid, haematopoietic and related tissue |
| Other cancers | Everything else in C00-97 except c34 and C81-C96 |
| Immunological disease | |
| HIV | B20 –B23 Human immunodeficiency virus [HIV] disease |
| Certain disorders involving the immune system | D80-D89 Certain disorders involving the immune system |
| Sickle cell disease | |
| SCD | D57 Sickle-cell disorders |
| Cystic fibrosis | E84 Cystic fibrosis |
| Organ transplantation other than kidney | |
